# Demonstration of four entities of appendicitis in China through studying cluster/outbreak

**DOI:** 10.1101/2020.02.24.20026021

**Authors:** Yi-Tian Guo, De-qiang Ye, Gui-fang Yang, Guozhen Liu, Xiao-chen Cui, Shi-Yun Tan, Yi Guo

## Abstract

**Objective:** To differentiate different entities of appendicitis through studying cluster/outbreak, ascertain common setting of cluster/outbreak, and provide the epidemiological evidences of infectious etiology of appendicitis.

**Background:** Differential diagnosis and management for perforated appendicitis and non-perforated appendicitis and the infectious etiology of appendicitis are current hot topics.

**Methods:** Field investigation for Tibetan students were carried out and reports published in English and Chinese medical journals were reviewed.

**Results:** The literature review included 473 patients in 7 cluster/outbreaks of appendicitis in 6 provinces and autonomous regions. All the clusters/outbreaks occurred in group living units. We found two classic entities of appendicitis with natural history from non-perforated appendicitis to perforated appendicitis and two entities of non-perforated appendicitis. In classic entities, one may represent majority of sporadic patients and the other may represent partial sporadic patients with obvious gastrointestinal manifestation. In entities of non-perforated appendicitis, one was identical to features of sporadic non-perforated appendicitis and the other one is identical to the following Tibetan students and associated with Fusobacterium.

The field investigation for 120 Tibetan students with appendicitis showed that the resected appendices exhibited diffuse or focal hemorrhages and infiltration by eosinophils and by lymphocytes. Most patients had normal body temperature, white blood cell count and neutrophil count. This is a new entity of appendicitis.

The clusters/outbreaks of appendicitis showed the features of infectious disease in epidemiology. The entity of perforated appendicitis was not found.

**Conclusion:** Studying cluster/outbreak is a good method to differentiate different entities of appendicitis and infectious etiology.

## INTRODUCTION

In more than 100 years, there has been no breakthrough in the etiology of appendicitis, and it is still unsure how many different entities of appendicitis exist. Natural history of acute appendicitis has classically been described to often progress from an non-perforated appendicitis to perforated appendicitis.^1-2^ Majority of the patients have elevated body temperature, total white blood cells counts and neutrophils in differential.^3^ A new hypothesis has been proposed that perforated appendicitis and non-perforated appendicitis may be different entities with different natural history.^4-8^ This has become modern classification.^9^ The infectious etiology, differential diagnosis and management for perforated appendicitis and non-perforated appendicitis are current hot topics.^10-26^

Since the appendix is connected to cecum which is exposed to the environment outside of human body, appendicitis can be caused by different etiologies. Therefore, appendicitis should not be only one entity, but a group of different entities. The histological examination of sporadic patients is targeted at a certain stage of different entities of appendicitis and the histological changes of appendicitis can not be seen continuously for same entity of appendicitis. It is therefore the classic description of natural history of appendicitis and the modern classification are deduced, not evidenced.

Cluster/outbreak often results from common cause, therefore almost every patient in cluster/outbreak belongs to the same entity and appeared in certain stage of same entity. Connecting each stage, we can know full natural histories of different entities and accordingly demonstrate whether or not perforated appendicitis and non-perforated appendicitis are different entities.

In 1984, a cluster of true appendicitis occurred in a town of 8,000 people in the United States.^31^ Thirteen patients of appendicitis occurred during a 3-month period. Regarding this clustering of appendicitis,the Centers for Disease Control (CDC) stated that the cluster offered a unique opportunity to identify possible risk factors and to search for precipitating infectious agents, and encouraged reporting such cluster to CDC.^32^ Since then, no typical cluster of true appendicitis has occurred until 1997. In 1997, we found a more severe cluster of appendicitis among students at a high school in China.^33^ As this type of appendicitis had unique pathologic findings, namely, hemorrhage in the lamina propria and hyperplastic lymphoid follicles, We tentatively named this condition “acute hemorrhagic appendicitis.” In 2012, Fusobacteria were also found in appendices of our clustering patients.^34^ Since beginning of 2005, we have been looking for new cluster/outbreak of appendicitis. We found that clusters/outbreaks occurred in many provinces of China and some of them were reported in English and some in Chinese medical journals. The aim of this study was to provide a new method to demonstrate different entities of appendicitis, causal association between microbiota and different entities of appendicitis and to improve modern understanding of sporadic patients. A second aim was to confirm common settings of outbreak/cluster of appendicitis and to provide guidance to find new clusters/outbreaks of appendicitis worldwide. A third aim is to provide the epidemiological evidences of infectious etiology of appendicitis.

## METHODS

### Epidemiological, Clinical and Pathological Investigation

Field investigation of cluster/outbreak was carried out according to Reingold AL’s methods^35^ and literature review was carried out to find the other cluster/outbreak.

### Field Investigation

Since 1985, Tibetan students were enrolled in the schools of 20 provinces and municipalities of China. We learned that incidence rates of appendicitis increased in many of these schools. From beginning of 2005, we contacted four schools in four provinces where Tibetan students were enrolled and cluster/outbreak of acute appendicitis occurred there for years. Finally we selected Tibetan students as study population at a high school in Nanchang, Jiangxi Province, China, because this high school and the hospital that treated student with appendicitis were willing to collaborate with this investigation.

We performed retrospective study of prevalence of acute appendicitis among Tibetan students from 2005, and since then carried out surveillance to July 2018. Since 2010, cluster/outbreak has not occurred because the school enrolled children of Han government officials sent to support Tibetan. These children did not live in countryside of Tibet and were not susceptible population, therefore we stop collecting data after 2010. The patients’ clinical, laboratory and histological examination information were collected on a standardized data collection form.

The study was approved by ethic board at participating hospital and Wuhan University School of Medicine, written informed consent was obtained from each patients and the subjects were interviewed since 2005.

To be included in this study, the following inclusion criteria have to be met: typical clinical manifestations of acute appendicitis and histologic features of acute appendicitis. All histologic slides from appendectomy were re-examined in Department of Pathology, Zhongnan Hospital of Wuhan University.

To examine if there are unique epidemiologic features for Tibetan students, we selected more than 7000 students of the native Han Nationality enrolled at the same high school as control population during same period.

### Definition of Cluster/Outbreak and Entities of Appendicitis

We included reports on cluster/outbreak of acute appendicitis according to CDC’s definition of cluster/outbreak.^36^ See supplementary 1.

According to classic classification and modern classification, three entities of appendicitis may be found through study of cluster/outbreak in theory:(1) classical Appendicitis: It shows continuum from non-perforated appendicitis and perforated appendicitis; Perforation rate should be close to 9.9% because hospital-based study showed this value is from 9.9% to 30%;^19^ (2) non-perforated appendicitis (acute simple appendicitis and acute phlegmonous appendicitis)^9^: Almost all patients or all patients in the outbreak had non-perforated appendicitis; (3) perforated appendicitis (including gangrenous appendicitis)^9^: Almost all patients or all patients in the outbreak had gangrenous appendicitis or perforated appendicitis.

### Investigation of Causes of Cluster in Tibetan students

The epidemiological data included person and time distributions of patients, exposure history and living habits, and environmental sanitation at this school, which were obtained by visiting the school and interviewing the patients and Tibetan students.

In August 2005, we interviewed 72 students to explore possible causes of the cluster, especially causes of higher incidence in female students. Among them, there were 28 patients who had acute appendicitis before interview. These students were asked to write their answers in the questionnaire. The questions included: (1) why there were more patients in female students; (2) what differences of living habits there were between female and male students; and between patients and normal students; (3) whether there were unclean or spoiled food intake, type of food intake, ingestion of water not boiled and dinner party before the onset of symptoms, habits of washing hands before meal and after bowel movement, contact history with patients, such as, living in same bedroom, meal with patients, caring patients.

According to the results, we offer advices for prevention.

### Analysis of Susceptibility in Tibetan Students

We compared proportion of the patients among female students enrolled in the first year, the second year, the third year, the fourth year and so forth to analyze susceptibility of Tibetan students.

### Infectious Chain and Estimation of Possible Incubation Period

The method see supplementary 2.

### The Other Cluster/Outbreak

From English and Chinese databases of medical literature (PubMed, Embase, CNKI, WanFang, VIP, CBM), we found clusters/outbreaks of appendicitis occurred in 5 provinces and autonomous regions in China.^37-43^ Among them, 7 clusters/outbreaks presented histological diagnosis or data of body temperature, white blood cell count (WBC), neutrophil percentage (NP, Considering that the change of neutrophils was indicated as a percentage of neutrophils in literature of cluster/outbreak, the change of neutophils in the patients of Tibetan students was also indicated as NP), which included 473 patients total.

### Statistical Analysis

Statistically significant differences in incidence rates between the male and female students were assessed using chi-square tests. A *P* value <0.05 was considered significant. SPSS 17.0 was used for statistical analysis.

## RESULTS

### Epidemiological Findings

Location and population: the school is located in an urban district of the City of Nanchang where Tibetan students were enrolled from 1985. Length of schooling was 4 years. Most of them came from Tibetan countryside. They board in the school. The living condition and the hygiene were good overall. See supplementary 3

Before 2000, there were clusters of infectious diseases, eg, tuberculosis, varicella, mumps, etc. From 1998, the incidence rate of appendicitis increased in Tibetan students. After 2000, the patients were admitted to the collaborating hospital. This study included 973 (474 male and 499 female) Tibetan students enrolled at this high school. Among them, there were 120 students suffered from acute appendicitis (102 females and 18 males) and 117 among them admitted to the collaborating hospital. Patients’ age ranged from 12 to 17 years. The mean age was 14.3 years. Person and time distribution of the patients were showed in table 1. There was a female preponderance (female 20.4%, 102 of 499; male 3.8%, 18 of 474; chi-square=62.280, *P*≤0.001). The incidence peak appeared from 2004 to 2006. In 2003, epidemic of severe acute respiratory syndrome (SARS) occurred in China. Because isolation of visitors, disinfection and personal protection were carried out at this school for prevention of epidemic of SARS, there was only 1 patient with acute appendicitis.

**Table 1.** Person and time distribution of the patients in Tibet students

| Year | Male |  | Female |  | Total |  |
| --- | --- | --- | --- | --- | --- | --- |
|  | Number of Students | Number of patients(%) | Number of Students | Number of patients(%) | Number of Students | Number of patients(%) |
| 2000 | 136 | 1(0.7) | 158 | 8 (5.1) | 294 | 9(3.1) |
| 2001 | 147 | 2(1.4) | 158 | 9 (5.7) | 305 | 11(3.6) |
| 2002 | 153 | 0(0.0) | 163 | 14 (8.6) | 316 | 14(4.4) |
| 2003 | 158 | 0(0.0) | 173 | 1 (0.6) | 331 | 1(0.3) |
| 2004 | 165 | 1(0.6) | 173 | 18(10.4) | 338 | 19(5.6) |
| 2005 | 176 | 3(1.7) | 168 | 19(11.3) | 344 | 22(6.4) |
| 2006 | 189 | 4(2.1) | 171 | 13 (7.6) | 360 | 17(4.7) |
| 2007 | 192 | 3(1.6) | 184 | 10 ( 5.4) | 376 | 13(3.5) |
| 2008 | 183 | 1(0.6) | 201 | 3 ( 1.5) | 384 | 4(1.0) |
| 2009 | 175 | 2(1.1) | 207 | 6 ( 2.9) | 382 | 8(2.1) |
| 2010 | 152 | 1(0.7) | 201 | 1 ( 0.5) | 353 | 2(0.6) |

The four year cumulative incidence rates in female students enrolled in 2001, 2002, 2003, 2004, 2005, 2006 were 26.8% (11of 41), 27.1% (13 of 48), 44.7% (21 of 47), 42.4% (14 of 33), 23.1% (9 of 39) and 19.3% (11 of 57) respectively before their graduation.

Many patients were classmates and/or roommates (data not shown). They had a contact history and suffered from acute appendicitis sequentially.

There were no obvious seasonal variations of patient occurrence among the Tibetan students. Cluster/outbreak of patients with acute appendicitis never occurred among more than 7000 native students at this school.

### Clinical and Laboratory Data in Tibetan Students

117 of the 120 patients operated on were from the collaborating hospital, from which clinical and laboratory data were obtained for the study; the other 3 patients, who were operated on but whose clinical and laboratory data were not obtained, were not hospitalized at the collaborating hospital.

The Clinical Features were presented in table 2. The patients’ clinical symptoms are similar to classic appendicitis, but patients’ results of body temperature and routine blood tests exhibited an obvious different pattern, namely, only a minority of the patients had mildly elevated body temperature and WBC (10.5 ×10^9^/L to 15 ×10^9^/L) or elevated neutrophil percentage (range 84% to 85%) on differential.

**Table 2.** Clinical Features of 117 Patients at the Collaborating Hospital.

| Variable | Value | no./no. with |
| --- | --- | --- |
| **Symptoms** |  |  |
| Gender |  |  |
| Male | 17 / 117 | (14.5) |
| Female | 102 / 117 | (87.2) |
| Fever | 12 / 117 | (10.3) |
| Nausea | 34 / 117 | (29.1) |
| Vomiting | 14 / 117 | (12.0) |
| Right lower quadrant pain (the initial abdominal symptom) | 80 / 117 | (68.4) |
| Midabdominal Pain migrating to the right lower quadrant (the initial symptom) | 43 / 117 | (36.8) |
| Tenderness in at or near McBurney's point. | 117 / 117 | (100.0) |
| Rebound tenderness | 58 / 117 | (49.6) |
| Involuntary muscle spasm. | 60 / 117 | (51.3) |
| Diarrhea | 0 / 117 | (0.0) |
| **Investigations** |  |  |
| White blood cell counts |  |  |
| Increased | 19 / 117 | (16.2) |
| Decreased | 7 / 117 | (6.0) |
| Neutrophil percentage |  |  |
| Increased | 12 / 117 | (10.3) |
| Decreased | 10 / 117 | (8.5) |
| **Histologic findings\*** |  |  |
| Hemorrhage in the lumen of the appendix | 77 / 116 | (66.4) |
| Diffuse or focal hemorrhage in the lamina propria or hyperplastic lymphoid follicles | 77 / 116 | (66.4) |
| Lymphocytic infiltration in epithelium | 67 / 116 | (57.8) |
| Infiltration of eosinophils | 86 / 116 | (74.1) |
| Lymphocytic infiltration in the serosa layer. | 50 / 116 | (43.1) |

\* Histologic slides from 116 patients were obtained

The mean length of time between onset of symptoms and operation (patients’ time) was about 4.7 days and the longest length of time between onset of symptoms and operation was 31days. Some patients had repeated onset of symptoms for several months.

During surgery, hyperemia and edema were observed on the appendix of each patient; however, no perforation was found and no purulent surface exudates were identified except for 7 patients. Microscopic stool examination was performed on 56 of 99 patients and no red blood cells were found. Platelet counts and prothrombin times were within the normal range in all patients.

### Histologic findings in Tibetan students

116 patients were examed pathologically. The resected appendices exhibited diffuse or focal hemorrhages in the lamina propria or within hyperplastic lymphoid follicles and in the lumen of the appendix of patients (Fig. 1A); Lymphocytic infiltration of the epithelium in the luminal surface (Fig. 1B) and serosa layer were also observed. there were infiltration of eosinophils in the lamina propria (Fig. 1C), submucosa, lymphoid follicles, and the muscle layers (Fig. 1D). Percentages of histologic findings were showed in table 2. Among the 116 patients, only 8 patients were diagnosed in collaborating hospital as acute phlegmonous appendicitis, 2 patients acute gangrenous appendicitis and the rest acute simple appendicitis according to classic histological diagnosis.

**Fig. 1.**
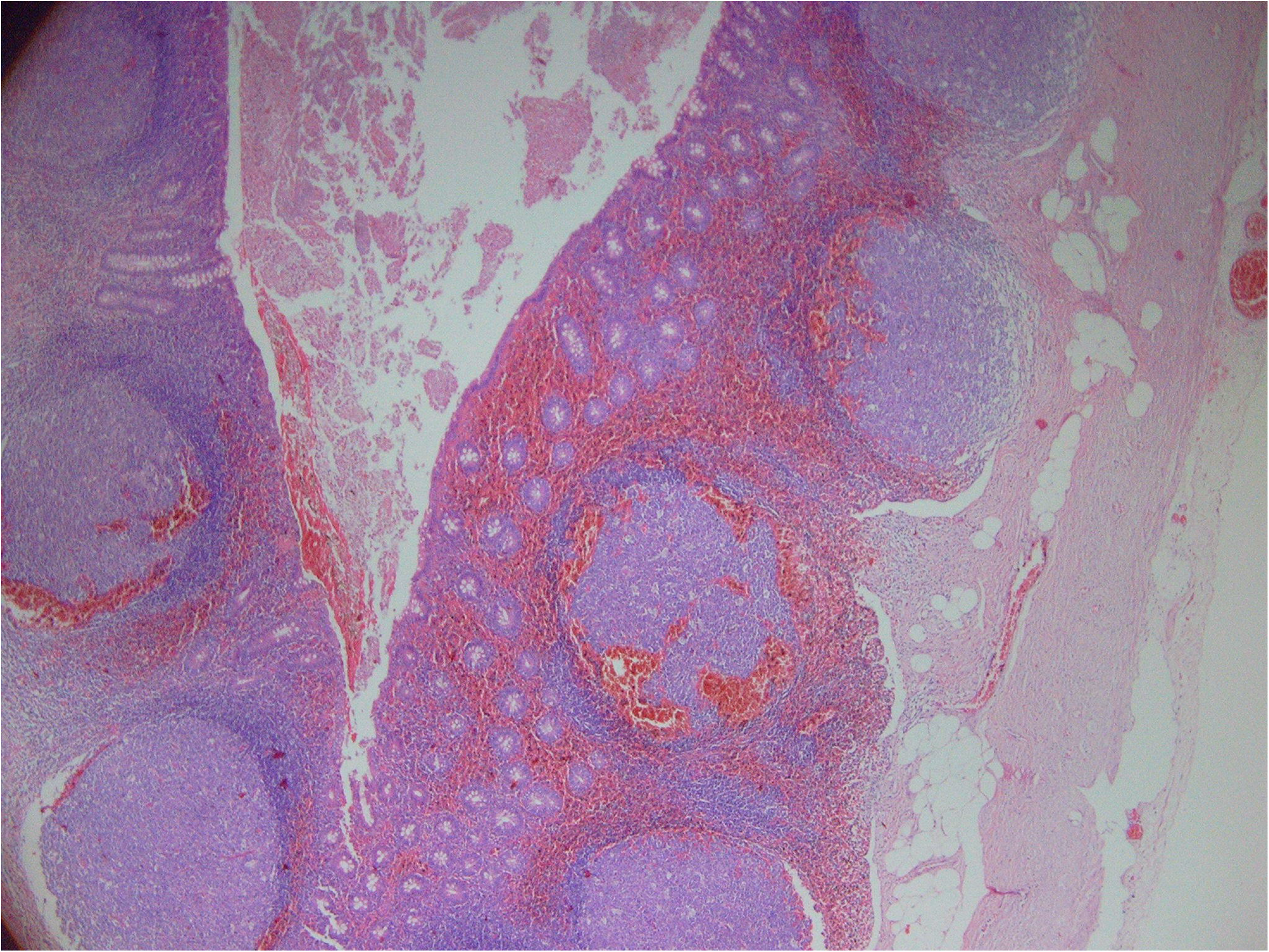
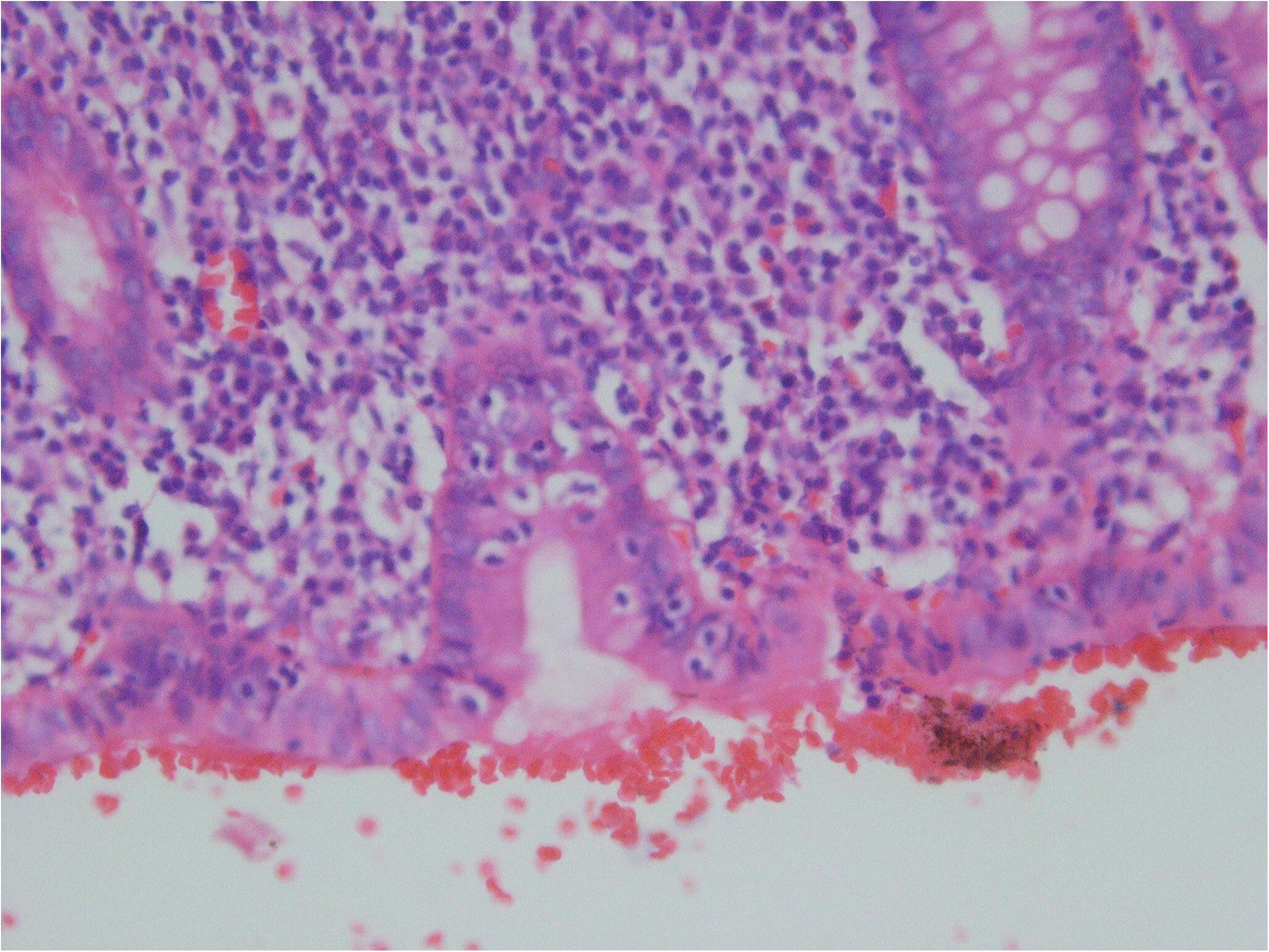
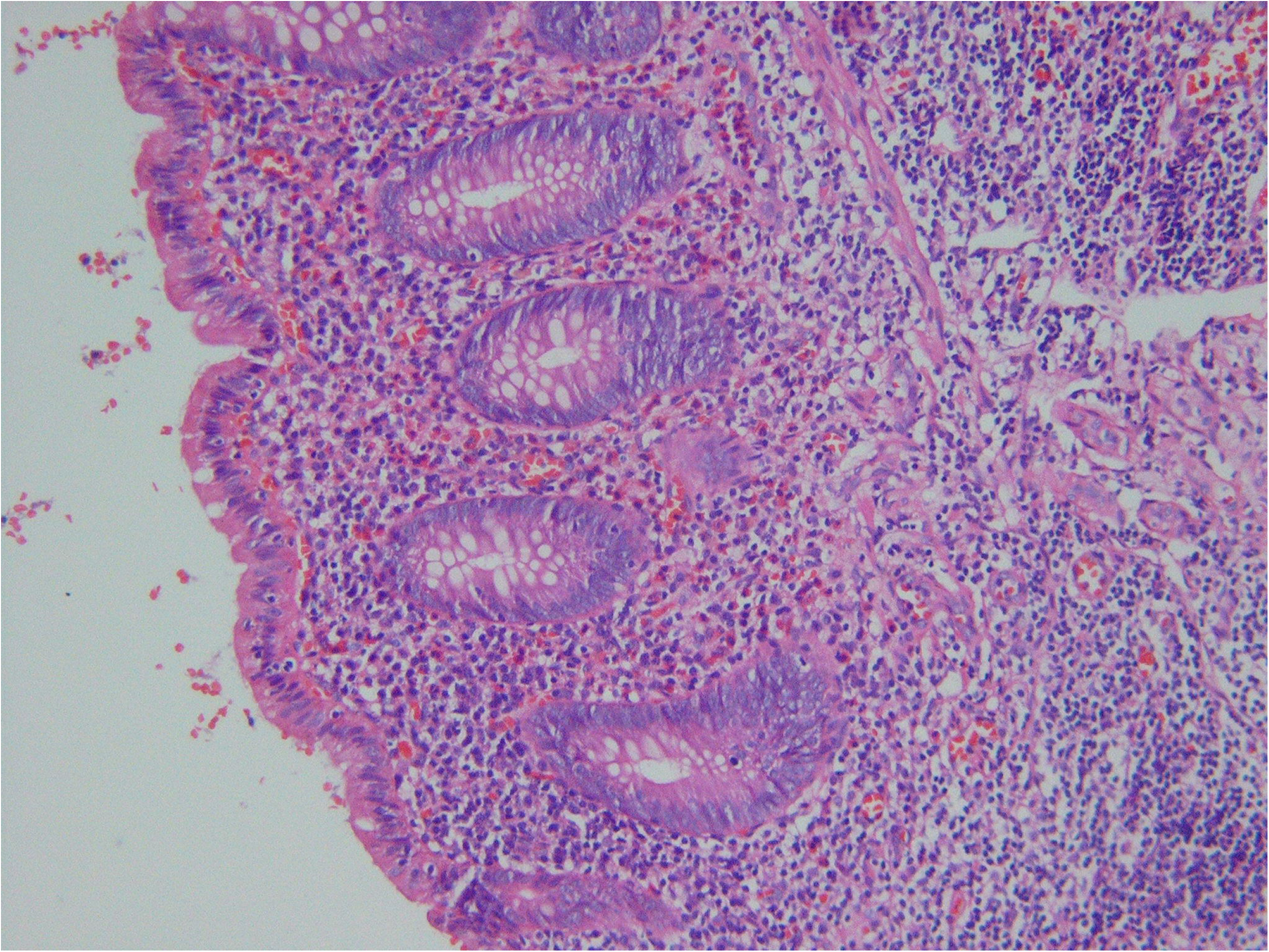
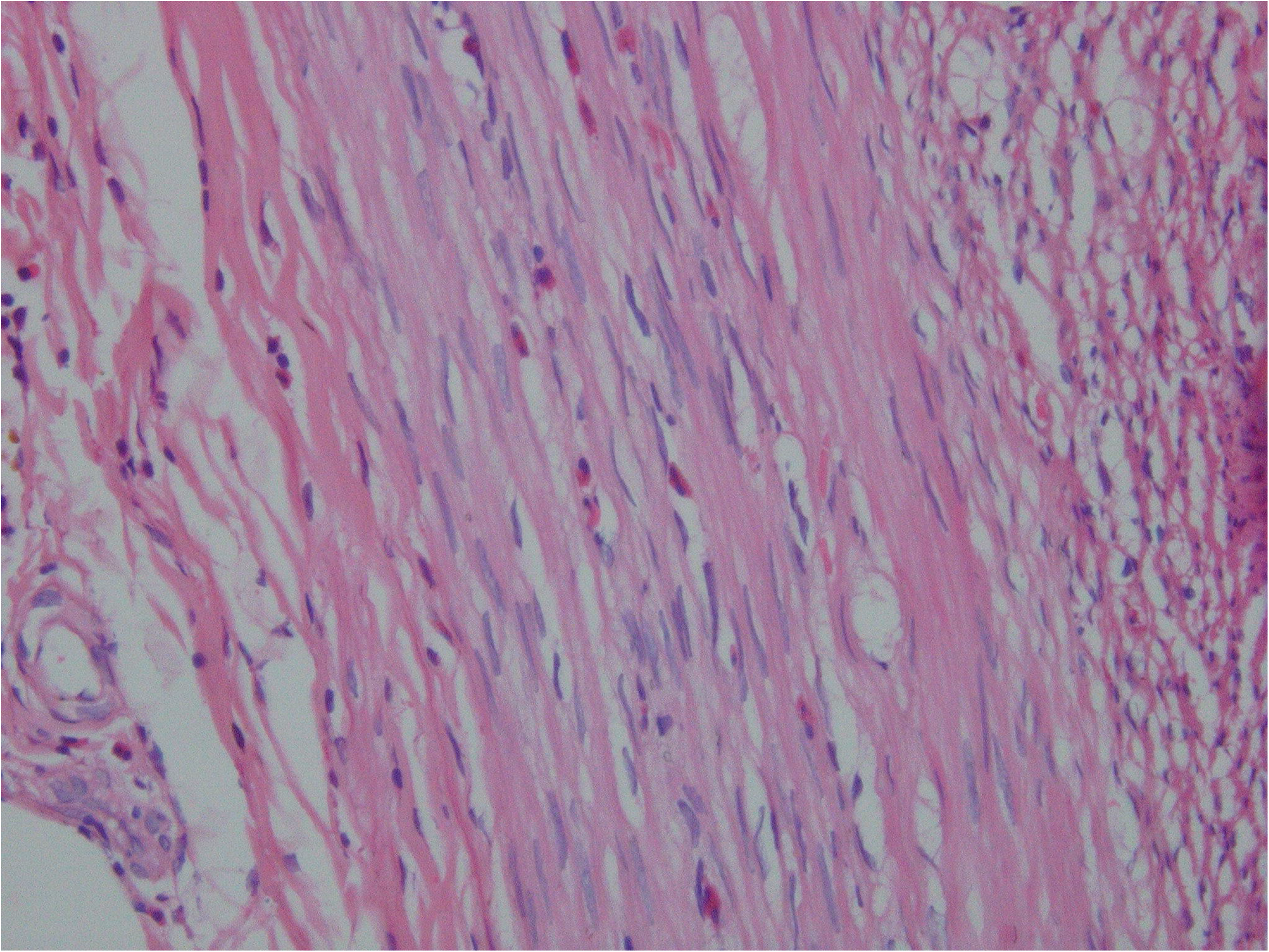
Histologic features of hemorrhagic appendicitis, as shown by hematoxylin and eosin stain. (**A**) diffuse hemorrhages in the lamina propria and lymphoid follicles as well as in the lumen of the appendix (original magnification, ×40). (**B**) The epithelium is infiltrated by lymphocytes (original magnification, ×400). (**C**) Infiltration of the lamina propria by scattered eosinophils, in addition to the lymphocytes and plasma cells. The epithelium is also infiltrated by lymphocytes (original magnification, ×200). (**D**) Eosinophilic transmural infiltration of the muscle layers (original magnification, ×200).

**Fig. 2.**
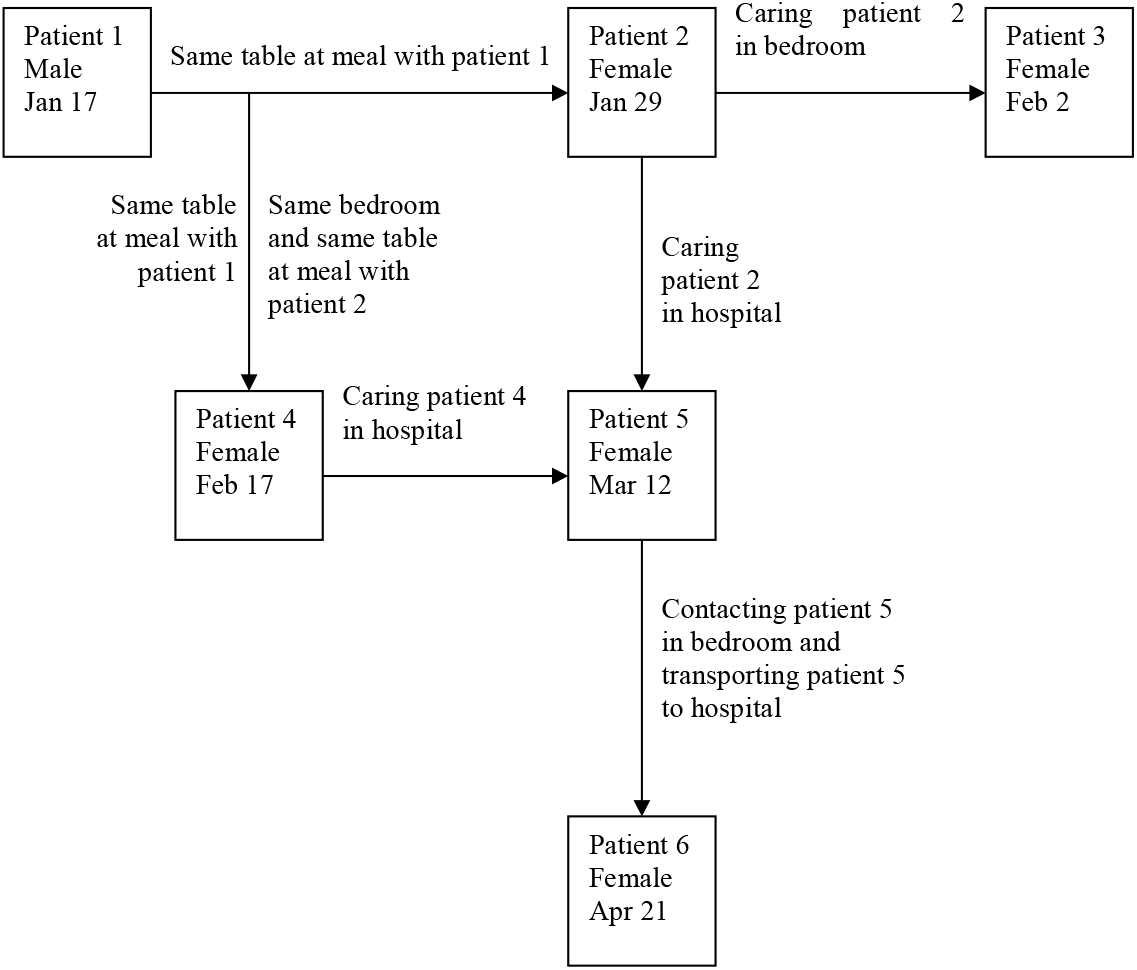
Infectious chain and temporal relation between contact and the onset of symptom of patients among Tibetan students who enrolled in Sep. 2006. Dates of onset of them are in 2007.

### Investigation of causes of cluster and measures for prevention

Most of 72 students we interviewed thought that the main difference of living habits between male and female students, and between patients and normal students were as follows: 1) Female students had a habits of staying in bedrooms, in contrast to male students who like outdoor activity; 2) Female students had a habit of eating snacks and sharing them with each other whereas male students did not have the habit; 3) The patients like staying in bedrooms and eating snacks and sharing them more than normal students; 4) Most patients had a contact history with their preceding cases before onsets of symptoms; 5) Toilets were built in bedrooms and most students do not have a habit of washing hands before meal and after bowel movement.

Contraposing the results, we offered the suggestion to carry out health education for students, especially for new students : 1) Washing hands before meal and after bowel movement; 2) Flushing and cleaning toilets after using toilet; 3) Escorting patients to the hospital and cleaning fomites once patients appeal; 4) Washing hand and clothes after contacting patients. 5) Limiting eating snacks.

Since August of 2005, incidence rates decreased year by year, table 1.

### Analysis of Susceptibility

See supplementary 4.

### Infectious Chain and Possible Incubation Period

See supplementary 5.

### The Other Cluster/Outbreak

See table 3. The epidemiological features of infectious diseases will be summarized in discussion. According to table 3, there are two entities of classic appendicitis (cluster/outbreak 1-2,^37^ 51 patients; cluster/outbreak 3^38^ and cluster/outbreak 6-7,^40-43^ Total 283 patients) and two entities of non-perforated appendicitis (cluster/outbreak 4,^31^ 31patients and cluster/outbreak 5,^39^ 108 patients). We did not find cluster/outbreak where most patients had gangrenous appendicitis or perforated appendicitis. In other words, we did not demonstrate perforated appendicitis as an independent entity.

**Table 3.** Epidemiologic, clinical and histological data from cluster/outbreak of appendicitis in China

| Studies | Settings | Epidemiological data | Clinical data | Histological data |
| --- | --- | --- | --- | --- |
| Xiao<sup>37</sup><br>1991<br>outbreak 1-2 | A middle school for outbreak 1 and a college for outbreak 2, Jilin city, Jilin Province | Total 51 patients occurred in outbreak 1 and outbreak 2. Outbreak 1: 36 students with acute appendicitis and 19 students with gastroenteritis after eating overnight food for breakfast in dining hall.<br>Outbreak 2: 15 students with acute appendicitis and 24 students with gastroenteritis after eating 24 hours left over food in dining hall.<br>Patients' ages: 16 ~ 20 years.<br>The authors did not present female number and male number of patients with appendicitis.<br>Each patient had nausea, vomiting and diarrhea.<br>Transmission route: food-borne transmission. | 76.5% (39 of 51) patients with acute appendicitis had body temperature of 37.3<sup>0</sup>C ~ 38.5<sup>0</sup>C; No bloody purulent stool and no tenesmus.<br>Incubation period : 3 h ~ 12 h after meals.<br>Average patients' time: not presented. | 46 patients had phlegmonous appendicitis; 5 patients catarrhal appendicitis; Gross examination: 26 had stink pus around appendix among these patients. |
| Yang<sup>38</sup><br>1991<br>outbreak 3 | University, Lanzhou City, Gansu Province | Since 1950, the incidence rate of appendicitis has been increased at Northwest Nationalities University for decades. Because the associated data was missing, the authors only collected data from 1981 to 1985. During this period, among 1730 Ethnic students enrolled, 59 had acute appendicitis:<br>Xinjiang Uyghur students: 4.6% (13/282 ) ; Tibetan students: 4.6% (12/263); Kazak students: 4.2% (3/72); Mongolian students: 3.8% (10/263); Hui students: 2.5% (13/527); the other ethnic students: 2.5% (8/323). However, native students' incidence rate for acute appendicitis is 0.2% (17/7100) at another university simultaneously in the same city (p<0.05).<br>Transmission route: not presented. | Proportion of patients in each class: Freshman: 50.8% (30 patients); Sophomore : 30.5% (18 patients); Juniors: 15.3% ( 9 patients); Seniors: 3.4% (2 patients).<br>It indicated that new students were more susceptible.<br>Patients' time: not presented. | 17 ethnic students had acute simple appendicitis. 23 phlegmonous appendicitis and 19 gangrenous appendicitis. Among these 19 patients, 11 had peritonitis caused by perforation. |
| Guo<sup>33</sup><br>2004<br>outbreak 4 | Middle school,<br>Wuhan City,<br>Hubei Province | Cluster phase: From April 10, 1997 to June 11, 1997, 11 patients occurred at a high school, with 3.8% (10 /290) students.<br>Post cluster: From the end of the initial cluster until June, 2000, 20 additional outbreaks were encountered.<br>Patients' ages: 16 ~ 20 years<br>More female patients (6.5%) than male patients (1.9%). Many patients had a history of mutual contact before the onset of symptoms.<br>Only a minority of the patients had mildly elevated body temperature and white blood cell counts or differential white blood cell counts<br>Pathologically, the resected appendices revealed hemorrhage with infiltration of lymphocytes and eosinophils.<br>All students were from remote area and isolated areas. New students were more susceptible.<br>Transmission route: indirect contact transmission.<br>Pathogen: Fusobacterium<sup>34</sup>. | Among total 29 patients operated on in collaborating hospital, 4 patients' body temperatures: more than 37°C (37.1°C, 37.3°C, 37.8°C, and 37.8°C). 6 patients' WBC: 11.3 ×10<sup>9</sup>/L ~ 12.5 ×10<sup>9</sup>/L. 12 patients' NP: 72% ~ 85%.<br>Average patients' time: about 50 hours. | Of 29 patients examined pathologically, 2 were diagnosed as phlegmonous appendicitis and 27 as acute simple appendicitis. Histological examination exhibited diffuse or focal hemorrhages and infiltration by eosinophils and by lymphocytes in the lamina propria or within hyperplastic lymphoid follicles. |
| Lu<sup>39</sup><br>2005<br>outbreak 5 | Camp,<br>Lanzhou City,<br>Gansu Province | During 8 week period of military drill from 1997 to 2001(Authors did not introduce month and date), 108 new soldiers (103 male and 5 female) with acute appendicitis.<br>Transmission route: not presented.<br>Patients' ages: 17 ~ 22 years. | Majority of patients had elevated WBC and elevated NP in differential WBC.<br>Patients' time: 3h ~ 72h.<br>Average patients' time: not presented. | Acute simple appendicitis and acute phlegmonous appendicitis. The authors did not report proportion of the two types of appendicitis. |
| Yan<sup>40-41</sup><br>2008<br>outbreak 6 | Camp,<br>Tibet | During period of training new recruit soldiers (from Dec. each year to March, the next year) from 2004 ~ 2007, 189(175 male and 14 female) with appendicitis; Ages: 16 years ~ 20 years. mean age: 18.8 years.<br>Transmission route: not presented. | 35 patients' body temperature: 36.5℃ ~ 37.2℃; 116 patients' temperature: 37.3℃ ~ 38.0℃; 38 patients' temperature > 38℃.<br>Patients' WBC :12×10<sup>9</sup> /L ~ 20 ×10<sup>9</sup> /L and 24 patients of them > 20 ×10<sup>9</sup> /L; Patients' NP: 72% ~ 93 %.<br>Patients' time: For most patients 2h ~ 72 h and for 15 more patients > 72 h.<br>Average patients' time: not presented | 97 patients had acute simple appendicitis; 68 patients phlegmonous appendicitis; 24 patients perforation. Among these patients, 2 periappendicular abscess. |
| Li<sup>42-43</sup><br>2008<br>outbreak 7 | Middle school,<br>Qingdao city,<br>Shandong Province | 660 Xinjiang Ethnic students were enrolled from 2002 to 2007; 35(10 male and 25 female) students with acute appendicitis; Age: 16 ~ 18 years. The incidence rate for Ethnic students each year is much higher than that for native students and total relative risk was 89.36 for 2002-2007.<br>Transmission route: not presented.<br>Incidence rate of appendicitis decreased after change of life style and health habit. | The patients' body temperature: 37.1℃ ~ 39.3℃. Patients' NP: 71% ~ 89 %.<br>Patients' WBC: 3.9×10<sup>9</sup>/L-13.7×10<sup>9</sup>/L<br>The authors did not provide the proportion of patients with elevated WBC.<br>Patients' time: 1 h ~ 11 h.<br>Average patients' time: 5h. | 11 patients had acute simple appendicitis; 15 patients phlegmonous appendicitis; 3 patients gangrenous appendicitis; 6 patients chronic appendicitis. |

Natural history and features of Clusters/outbreaks 3 and cluster/outbreak 6-7 were identical to classic description of natural history from non-perforated appendicitis to perforated appendicitis. The different forms of inflammation in appendices for classic appendicitis were histological features of different stage of same entity, not different entity. The clinical features showed elevated body temperature, WBC and NC in majority of patients, which is similar to most sporadic appendicitis and suggest that the patients in clusters/outbreaks 3 and cluster/outbreak 6-7 were likely the same type of appendicitis as majority of sporadic patients.

In cluster/outbreak 1-2, the onset of patients occurred between 3 and 12 hours after eating overnight food. 90.2% (46/51) of patients had phlegmonous appendicitis and 76.5 % (39/51) had elevated body temperature. 51% (26 of 51) of patients had more stink pus around appendix which may develop to gangrenous appendicitis and perforation if surgery was not carried out. We classify outbreak 1-2 into classic appendicitis. Each patient had obvious digestive manifestation and occasional diarrhea^5, 23^. The patients had no bloody purulent stool and no tenesmus, therefore dysentery can be excluded. Cluster/outbreak 1-2 is different entities from clusters/outbreaks 3 and cluster/outbreak 6-7, and may represented partial sporadic patients.

Outbreak 4 and Tibetan students should belong to the new type of non-perforated appendicitis because they had the same epidemiological, clinical and histological features as Tibetan students. Most patients had normal body temperature, WBC and NC, and unique histological features with hemorrhage and infiltration by eosinophils.

Outbreak 5 also belongs to non-perforated appendicitis and its patients have elevated WBC and NC. Their histological and clinical features were similar to sporadic non-perforated appendicitis and may represent sporadic non-perforated appendicitis.

## DISCUSSION

We have presented 8 outbreaks of appendicitis occurring in 6 provinces and autonomous regions in China. As far as we know, this is the most detailed summarization of clusters/outbreaks of appendicitis. All clusters/outbreaks of appendicitis occurred in group living units. The features of distribution will provide methods to find new cluster/outbreak. Because appendicitis is not endemic disease, cluster/outbreak of appendicitis should also occur widely worldwide and can be found using same methods as we did in China. In fact, cluster/outbreak of appendicitis occurred more frequently than we realized in this paper. For example, we did not report 3 other schools where cluster/outbreak of appendicitis occurred in Tibetan students, because these schools were not willing to collaborate with us.

Through field investigation and literature review, we found two classic entities of appendicitis and two non-perforated appendicitis including a new entity of appendicitis. Cluster/outbreak 3 and cluster/outbreak 6-7 may represent majority of sporadic patients with classic appendicitis because their natural history, historical and clinical features were identical to that of the sporadic patients. Cluster/outbreak1-2, may represent partial sporadic patients with obvious gastrointestinal manifestations. The cluster of appendicitis occurring in the United States was also caused by food-borne transmission and belongs to classic appendicitis because perforation occurred in 31 percent of patients^32^, which indicate that appendicitis caused by food-borne transmission occurred worldwide.

Cluster/outbreak 5 was identical to features of sporadic non-perforated appendicitis and may represent majority of patients with this disease.

Tibetan students and patients in cluster/outbreak 4 represented a new entity of appendicitis. Because they had the following features: (1) Pathologically, the resected appendices revealed hemorrhage with infiltration by lymphocytes and eosinophils. The features can be distinguished from that of appendicitis resulting from the known infectious agents.^44^ (2) In clinical data, only a minority of the patients had mildly elevated body temperature and WBC counts or NC. (3) In epidemiology, there were the occurrence of clustering; Female patients were more common than male patients and they had a history of mutual contact before the onset of symptoms; New students had higher susceptibility. Cluster can be controlled through isolation of visitors and disinfection as evidenced in 2003 for prevention of SARS among Tibetan students. Considering that histological features of Tibetan students and cluster/outbreak 4 revealed not only hemorrhage in appendiceal tissues but also infiltration by eosinphils and so forth, we tentatively re-named “acute hemorrhagic appendicitis^33^” to “acute hemorrhagic and eosinophil-infiltrated appendicitis.” Although there are 2 patients with gangrenous appendicitis in Tibetan students, the incidence rate is very low. The reason is during 10-year period, partial patients may have classic appendicitis.

Compared with cluster 4, the cluster among Tibetan students lasted much longer and showed much higher incidence rates. We are surprised to find out that incidence rate of the female students enrolled in 2003 and 2004 who had surgical operations because appendicitis were 44.7% (21 of 47) and 42.4% (14 of 33) respectively. This may be since Tibetan students were from more remote and isolated areas, they were more susceptible in their new environment.

The patients’ time of this new type of appendicitis was much longer (4.7 day on average for Tibetan students) than that of sporadic appendicitis. Some study showed that patients’ time of 12, 24 and 36 hours increased relative risk for perforated appendicitis^23^. Our study suggests that it is not reliable to early diagnose perforated appendicitis and non-perforated appendicitis just through patients’ time.

Our results provided more sufficient epidemiological evidence to support infectious etiology of appendicitis. The reasons are as follows: According to living habits of Tibetan female students and analysis of history of mutual contact, we postulated that Tibetan students’ appendicitis resulted from indirect contact transmission; Finding of Fusobacterium nucleatum/necrophorum in cluster/outbreak 4 suggested that our infectious hypothesis is reasonable;^34^ Incidence rate of appendicitis decreased after intervention of life style and health habit; Analysis of susceptibility indicated that new students have higher susceptibility.

Like Tibetan students, the other cluster/outbreak also showed features of infectious disease in epidemiology. The students and soldiers from remote areas, the minorities, and female students and the new comers are more susceptible than native students and soldiers.^31,39-43^ High attack rates in female students were associated with their living habit^33^. New endemic location can form and persist for years and decades.^38,40-41, 42-43^ Transmission routes were associated with food borne transmission^37, 39^ and indirect contact transmission;^33^ Incidence rate of appendicitis lower after intervention of life style and health habit.^43^

Many gastrointestinal diseases were reported to mimic symptoms of appendicitis. In several outbreaks induced by enteric pathogens, patients who had appendectomies were found to have normal appendices, ^45-48^ but our patients from cluster/outbreak had true appendicitis demonstrated by histological examination.

The discovery and confirmation of new diseases need to be representative and reproducible^49^. Except cluster/outbreak 5, the other cluster/outbreak occurred at least twice and lasted for many years. All 593 patients in cluster/outbreak included various histological features and representative. Our study meets the above requirement.

Since 2009, new studies have provided evidence of association between microbiota and appendicitis.^10-14.50^ Through studying patients in cluster/outbreak, the difference in microbiota between different entities of appendicitis or different stage of same entity may be confirmed. Further we may make etiological diagnosis for appendicitis and confirm proportion of communicable appendicitis and non-communicable appendicitis clinically, and in future, improve diagnosis and treatment of appendicitis.

## Limitation

Some reports did not describe epidemiological features in detail^37-43^ or did not provide data of elevated body temperature,^38, 39^ WBC,^37-38, 42-43^ NC^37-38^ and/or average patient’s time.^37, 38, 39, 40, 41^ Estimation of incubation period is crude.

## Conclusion

We demonstrated that cluster/outbreak of appendicitis is not unusual. There may be four entities of appendicitis including a new entity, which reveals the features of infectious diseases in epidemiology. The study of cluster/outbreak appendicitis can correct the bias caused by sporadic cases.

## Data Availability

I can prodide all data referred to in the manuscript.

## Conflict of interest

No conflict of interest was declared.

## Notes

### Competing Interest Statement

The authors have declared no competing interest.

### Funding Statement

No funding

